# Smart Wristband Monitoring: A Caregiver-Oriented Mobile Application for Tracking Cognitive Decline in Mild Cognitive Impairment Patients

**DOI:** 10.1101/2024.05.31.24307874

**Authors:** Barış Ceyhan, Semai Bek, Tuğba Önal-Süzek

## Abstract

Mild Cognitive Impairment (MCI) is the transitional phase between the typical expected memory problems related to age and the more serious decline of dementia. Therefore, monitoring progress from MCI to dementia is critical to slowing down this cognitive deterioration. Research shows that proper nutrition, routine brain, and physical exercise accompanied by proper care slow down the disease significantly. However, most of the patients do not perform these exercises regularly and their vital health data is not monitored properly, leaving patients most of the time with only basic care.

In this study, for the first time to our knowledge, we aimed to develop a mobile application that will enable caregivers to continuously monitor the vital health, medication, activity, and location of the patients with MCI with a smart wristband while enabling them caregivers to track the progress of the disease by a machine learning model that tracks MMSE of the patient using speech. Based on our research so far, this caregiver-oriented approach with the ability to track progress of the disease using the convenience of a mobile application is a unique attempt in the field. Patient profile along with collected data is correlated with tracked MMSE scores of the patients to come up with recommendations and findings about the patients. Caregivers are relieved to be notified about critical health data thresholds, progress, and condition of their patients.

## Introduction

Dementia ranks seventh among the leading causes of death and stands out as a significant contributor to disability and dependency among older individuals worldwide [1]. While age is a critical risk factor in dementia; being physically active, not smoking, controlling weight, a healthy diet and maintaining healthy blood pressure, cholesterol and blood sugar levels reduce this risk significantly [2].

There are studies that research correlation between vital health data and dementia progress. A study found out that older adults with Resting High Rate(RHR) >= 80 had a 55% increased risk for developing dementia [3]. Another study found sleep problems and duration are indicators of quick progress to dementia from MCI [4]. A study conducted with 46 adults with dementia demonstrated a direct correlation between MMSE results and walking rates [5]. In a recent systematic review of several medication adherence studies of dementia patients, nonadherence correlated with higher hospitalization or mortality rates, with the highest adherence rate observed at only 42%; however, telehealth home monitoring and treatment modification were identified as the only interventions to improve medication adherence [6].

Several fitness, location tracking applications are available on the market however there is not an application in the market that is caregiver oriented and can track the progress of dementia.

The focus of this paper and the developed application is to analyze the significance of vital health data in MCI patients, validated with a speech-based MMSE prediction test that can be conveniently conducted by a mobile application to track progress by caregivers. The novelty of our work in this study is that it is the first smart wristband based dementia monitoring software system according to our knowledge which aims to valuate the significance of health data parameters for tracking cognitive status decline rate in MCI patients.

## Methods

### Dataset

A three-month-long collection of vital health data was conducted on 30 participants aged over 60 between September 2023 and March 2024 per patient availabilities. All participants were equipped with the same smart fitness wristbands (Xiaomi MI Band 7) for the duration of the study. Summary of clinical variables of patients along with their MMSE scores performed within a month before the start of the data collection are displayed in Table 1. Ethics committee approval issued by Mugla University is supplied as translated from Turkish in S1 File. Patient consent form was collected in written format and translated form is supplied in S2 File. A mobile application has been developed that caregivers can register their patients, wristbands and allow them to monitor their health data along with their location. medication and activities while synchronizing data anonymously to the application database.

**Table 1.** Summary of the clinical variables of patients analyzed in the study.

| **n=30** |  | **Age range at the**<br>**start of the study**<br>**(60-69)** | **Age range at the**<br>**start of the**<br>**study (> 70)** |
| --- | --- | --- | --- |
| **Female** | MMSE < 20 | 6 | 7 |
| **Female** | MMSE > 24 | 1 | 2 |
| **Female** | MMSE 20-24 | 0 | 3 |
| **Total:** | 20 |  |  |
| **Male** | MMSE < 20 | 1 | 2 |
| **Male** | MMSE > 24 | 2 | 1 |
| **Male** | MMSE 20-24 | 2 | 2 |
| **Total:** | 10 |  |  |

### Data Collection

Existing tests aimed at diagnosing neurodegenerative diseases often struggle to effectively identify deviations from the typical cognitive decline trajectory during the initial stages of the disease. Particularly for patients with MCI, the benefits of passive data collection through devices such as smart wristbands—such as increased frequency of data acquisition, objectivity, and reduced patient burden—result in higher adherence rates and greater precision compared to active data collection methods [7]. While mobile phones do track some health data like steps, calories; patients older and with dementia symptoms tend to not carry their mobile phones continuously everywhere, but also the phones do not track health data like heart rate (HR), resting heart rate (RHR) and sleep patterns. While some phones are capable of tracking additional health metrics such as blood pressure and include GPS functionality [8], the data collection in this study utilized relatively affordable and straightforward Xiaomi MI Band 7 wristbands. These wristbands track only heart rate, resting heart rate, sleep patterns, steps, and active minutes due to their reasonable price, lightweight design, and ease of wear. In a research conducted for adults over 50 years of age, while participants generally found it stressful to use smart wristband technology, they found using such wristbands helpful due to their heart rate and movement tracking capabilities [9].

In our study, we deployed a mobile application that continuously synchronizes data from the wristband to application servers using Google Fit services, which seamlessly integrate with the device software. Using Google Fit as the proxy application enabled us to use any other smart watch or wristband that can synchronize with Google Fit in the market since most watches support Google Fit by default [10]. Google Fit platform allows software developers to build applications and services that can access fitness data from wearable devices, fitness apps, and health-related sensors. Developers can integrate Google Fit into their apps to access and manage user’s health and fitness data, including activity tracking, heart rate, sleep patterns, and more. This API enables developers to create innovative health and fitness-related applications while leveraging Google’s infrastructure and data sources. Google fit works with more than 30 smart watches and wristbands on the market, simply summarizes the data and provides the data to consumers with REST API endpoints. Just as it receives health data from the phone as an application, it also receives data from many auxiliary applications and provides access to them both from the application and through the Google Fit services. We used Google Fit API’s functionality to collect and synchronize data from both the phone and the smart wristband in one place. This makes it possible for the patient’s data to still be collected when he or she is not actively using the phone or the smart wristband. A limitation of using the Google Fit was the installation steps needed in addition to installing the mobile application. The requirement to have a Google Mail account discouraged some older users from installing the application. Moreover, whenever older patients needed to restart their phones for some reason, Google Fit did not start automatically in the background, and this caused delays in data collection. For future work we plan on adapting our software to the native API of the smart wristband for better patient cooperation causing less anxiety to older patients.

The cross-platform mobile application was developed with the assumption that caregivers will install the application to the patient’s mobile phone once and track their patients from their phone without the need of patient input. The additional software functionality added for caregivers like medication and location tracking, scheduling activities, and notifications for critical thresholds on health data, mobility and medication were added to encourage more patients into the study. For the accurate collection of medications and their Anatomical Therapeutic Chemical (ATC) codes, we have developed a scheduled task and a web service to update the latest medication names and ATC codes from the Turkish Ministry of Health weekly. Google Firebase Cloud Messaging has been implemented for notification services for reminders.

## Results

In our analysis we examined the smart wristband based categorical and non-categorical health data collected from 30 participants over a three-month period. The statistical summary in Table 2 outlines the distribution of various health variables, while Figures 3, 4, and 5 illustrate the relationships between cognitive scores and health metrics through PCA, correlation coefficient analysis, and chi-square analysis, respectively. Additionally, the integration of our previously published machine learning model allowed us to predict the MMSE scores, revealing insights into the impact of physical activity on cognitive function and overall well-being.

**Figure 1.**
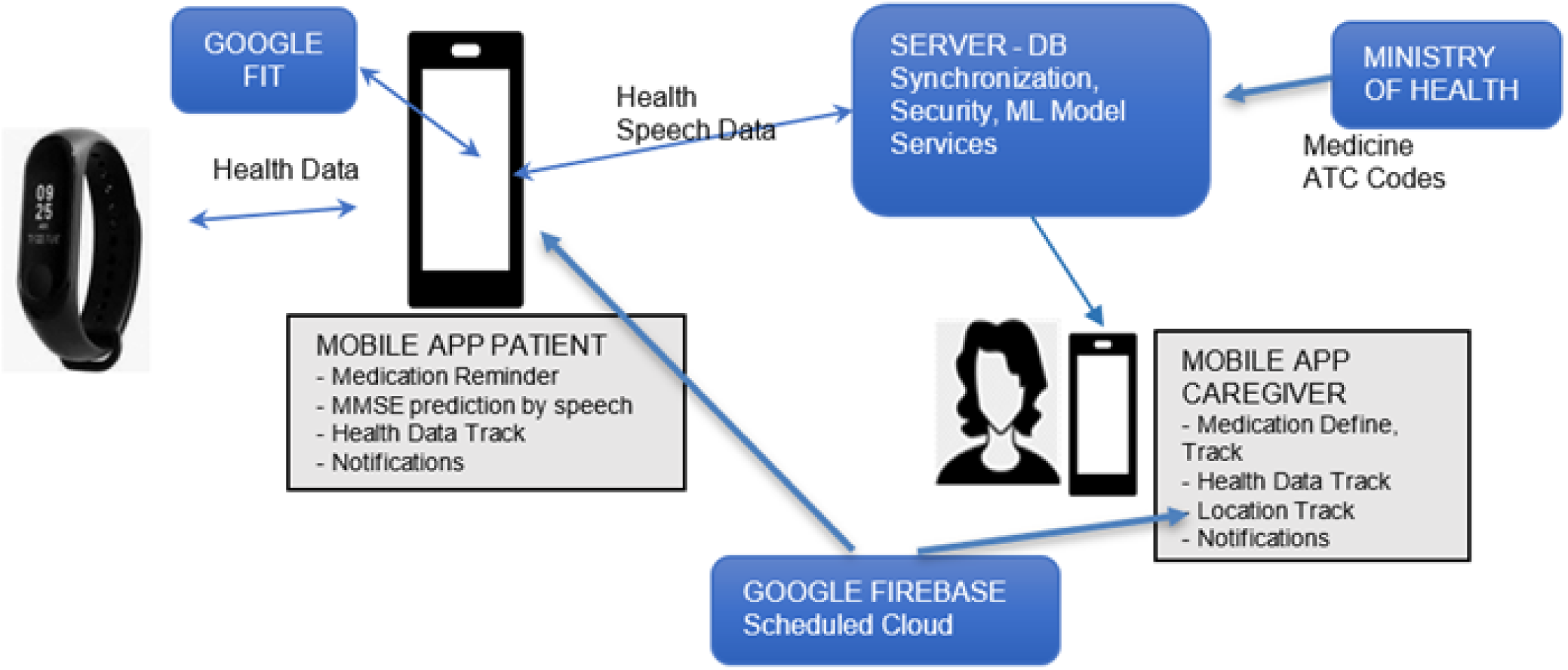
Application component diagram.

**Figure 2.**
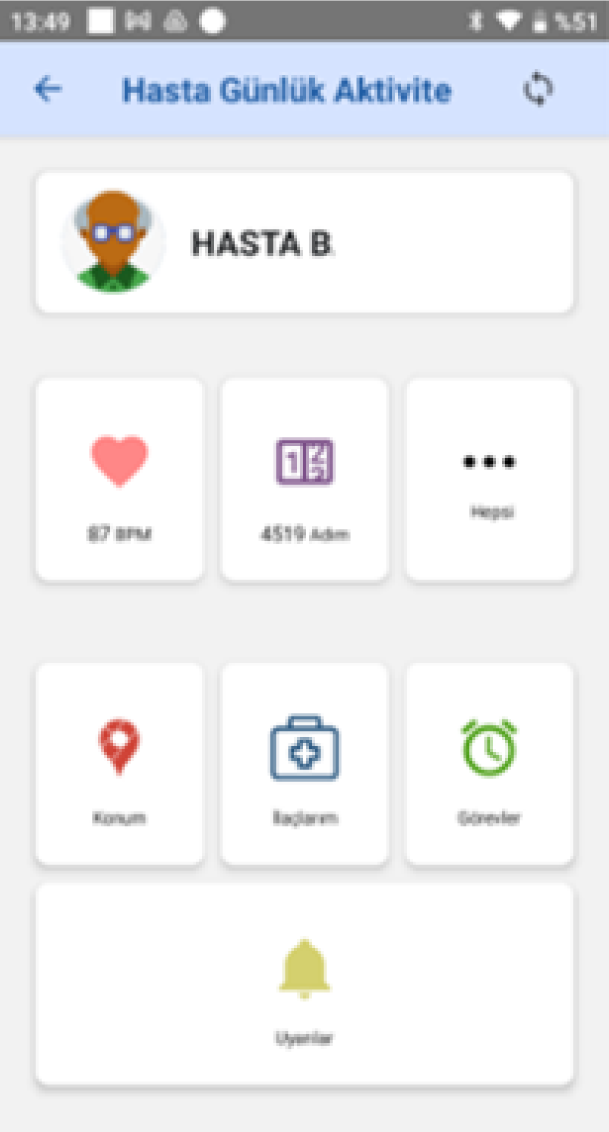
Mobile application front-end example health data summary.

**Figure 3.**
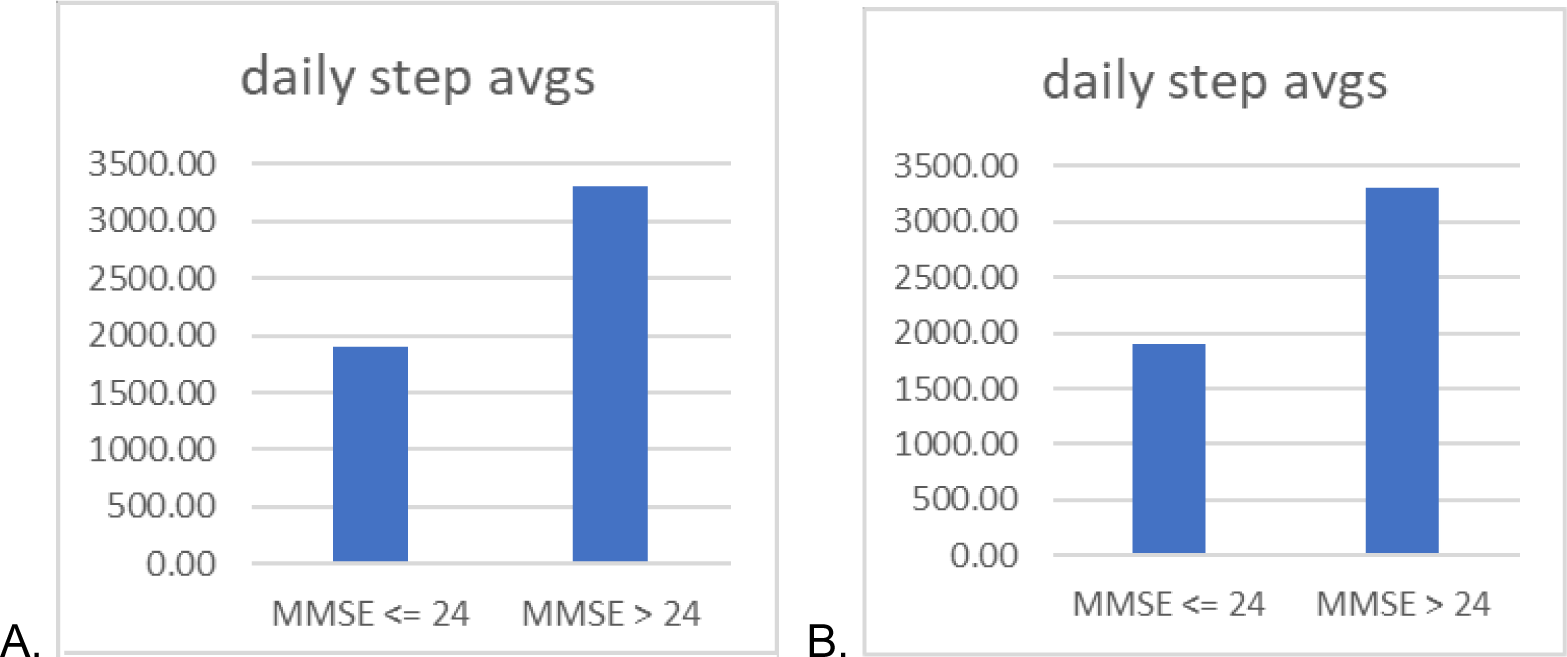
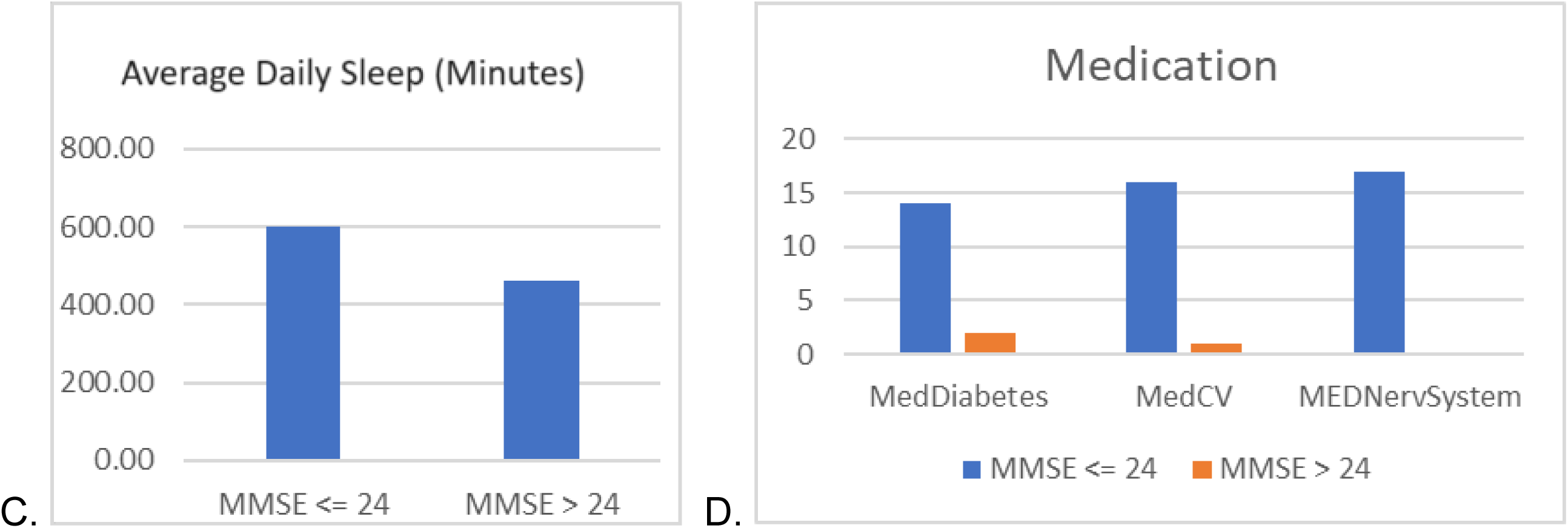
(A) Aggregated statistics on average daily sleep (B) HR and RHR (C) daily sleep in minutes (D) number of medications taken by particapants.

**Figure 4.**
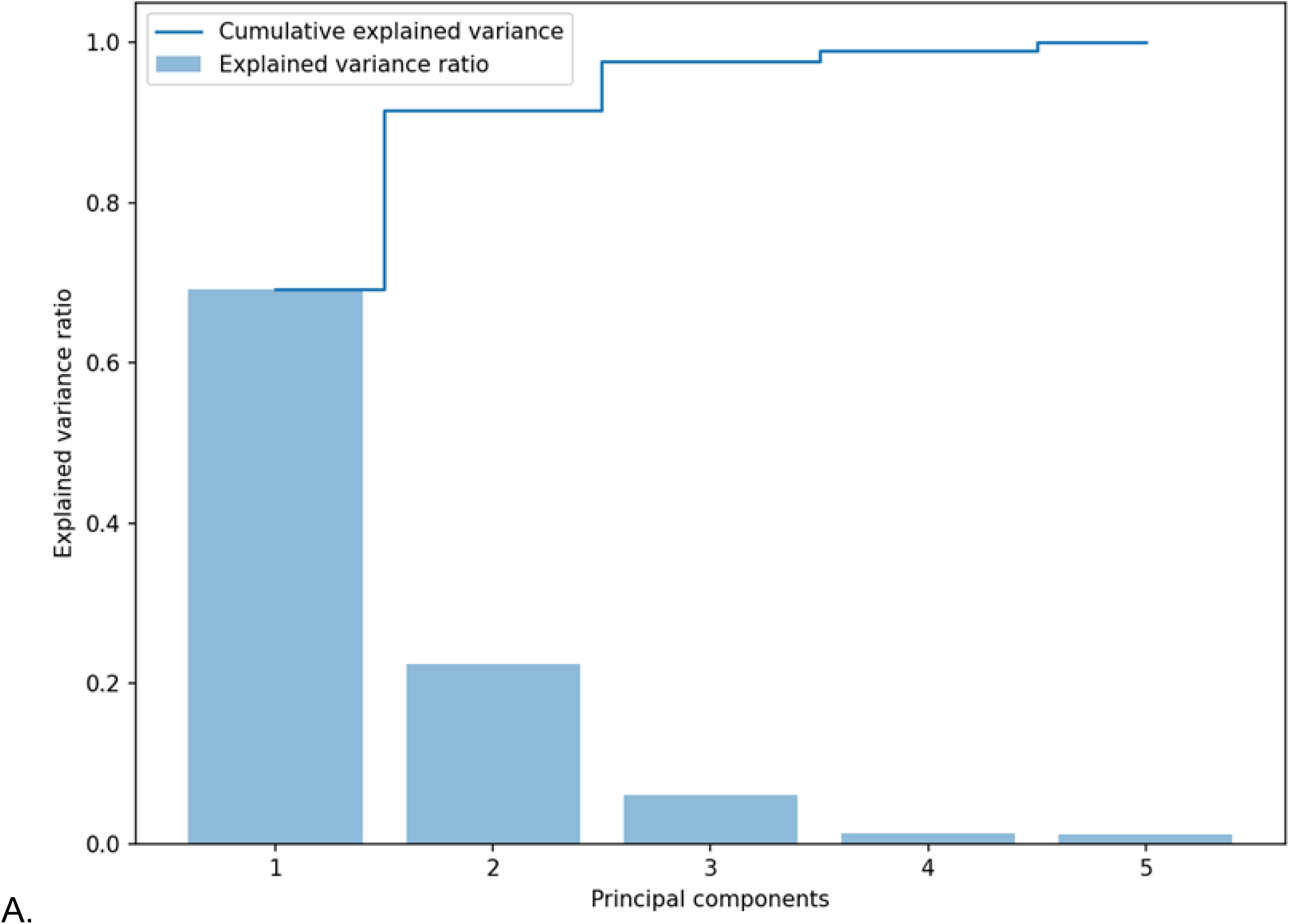
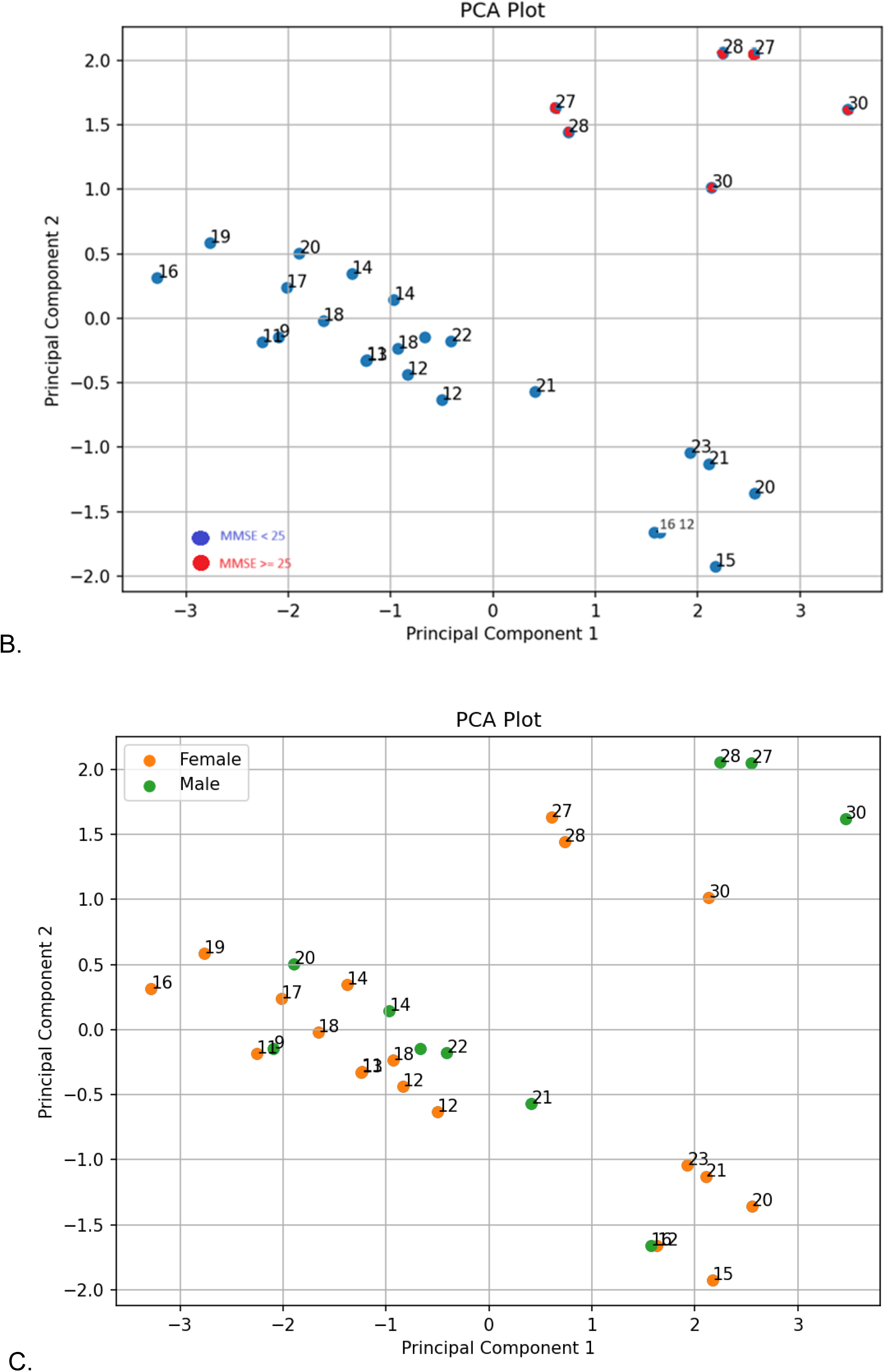
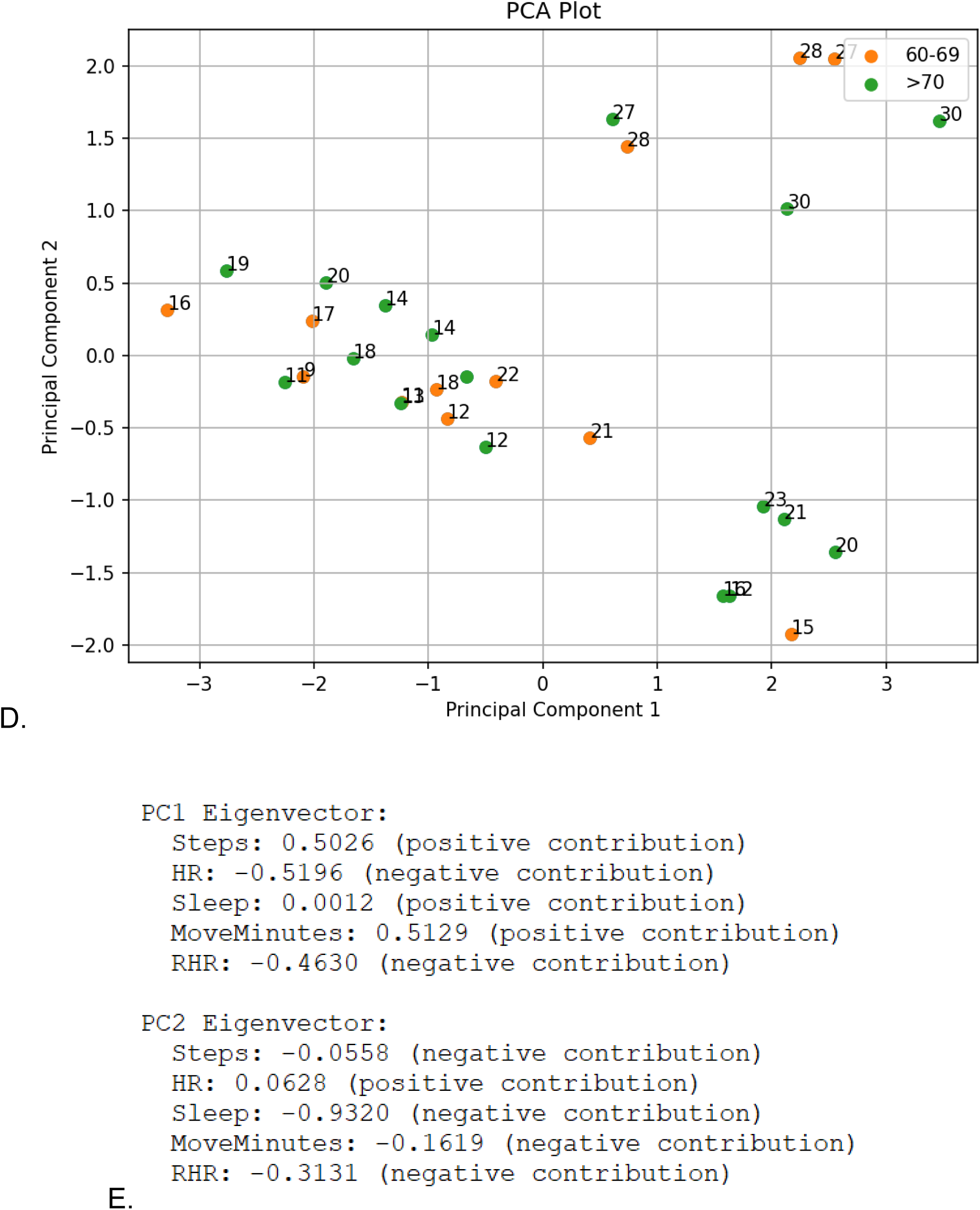
(A) Cumulative Explained Variance and Explained Variance Ratio (B) Top two dimensions of the PCA analysis based on the smart wristband health data variables of patients show the spread between the patients with higher cognitive scores (MMSE >= 25) cluster together and away from the patients with lower cognitive scores (MMSE < 25) (C) Gender distribution in top two dimensions (D) Age distribution in top two dimensions (E) Eigenvectors of the first five principal components show that sleep is the highest negative contributor and Steps and Move minutes are the highest positive contributor to the cognitive test.

**Figure 5.**
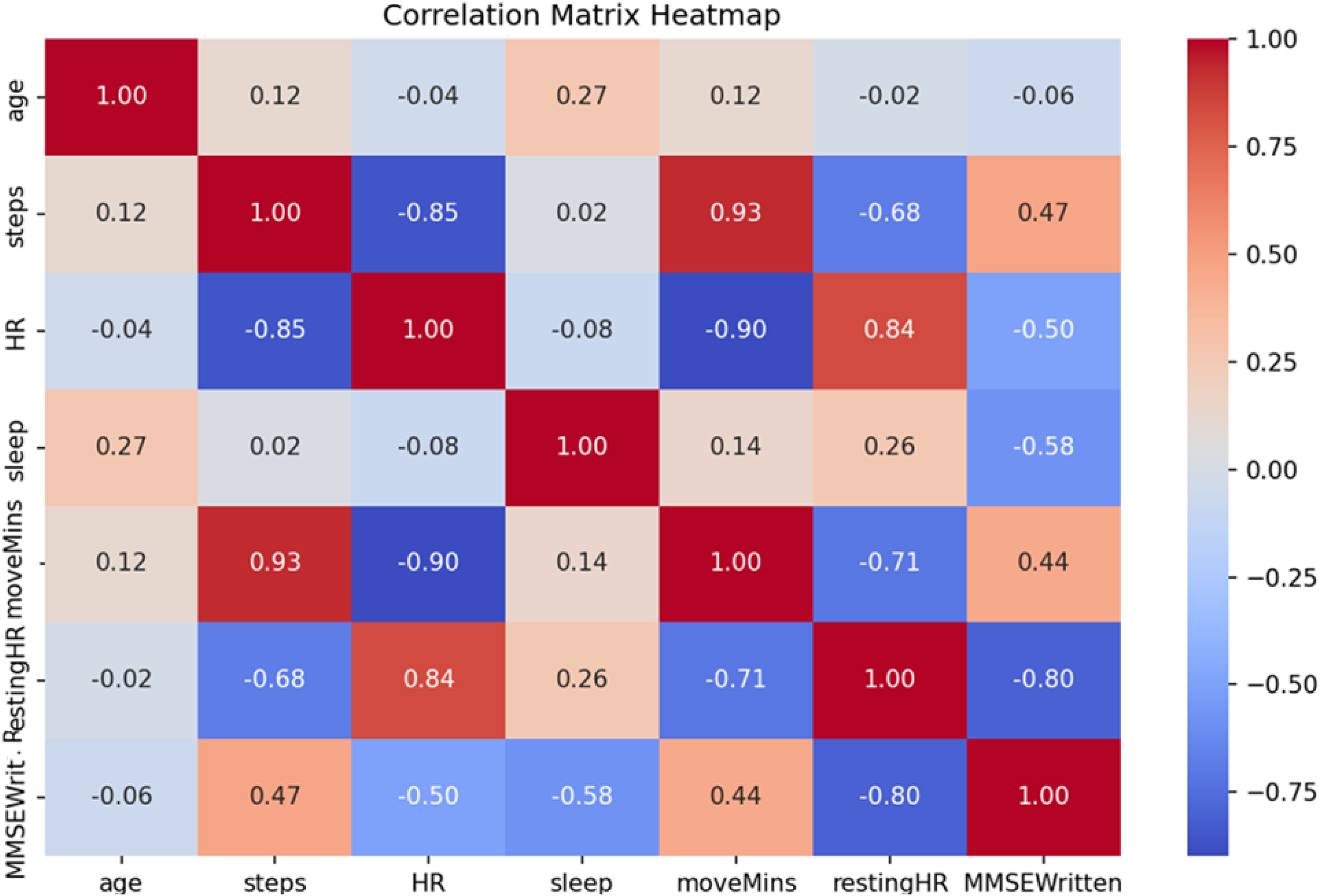
Correlation coefficient heatmap of all continuous variables show that number of steps and number of minutes of movement are significantly positively correlated with higher MMSE score with p-value ≤ 0.05. Resting HR Rate is negatively correlated with higher MMSE score with p-value of 0.007.

**Table 2.** Statistical summary of the smart wristband based health data variables of the 30 patients.

|  | Mean | std | min | 25% | 50% | 75% | max |
| --- | --- | --- | --- | --- | --- | --- | --- |
| age | 70.69 | 4.83 | 63.00 | 67.00 | 71.00 | 74.00 | 78.00 |
| gender |  |  |  |  |  |  |  |
| steps | 2195.66 | 1119.54 | 679.83 | 1321.36 | 1763.12 | 2903.48 | 4379.49 |
| HR | 70.87 | 7.11 | 59.06 | 64.82 | 71.08 | 76.32 | 82.87 |
| **Sleep**<br>**(mins)** | 572.69 | 73.11 | 422.65 | 533.69 | 572.52 | 617.98 | 682.23 |
| **Move**<br>**Minutes** | 53.45 | 27.77 | 13.58 | 32.93 | 44.55 | 83.92 | 100.53 |
| **Resting**<br>**HR** | 66.54 | 5.71 | 56.84 | 61.05 | 67.68 | 70.33 | 78.18 |

Figure 3 A, B and C show aggregated health data classified based on MMSE scores of 30 Patients from the 3 months of collected data. In another study in literature with an average age of 73, a MMSE score of 24 was identified as the threshold indicating severe cognitive impairment [11] therefore we clustered each patient into high- and low-cognitive score groups based on the MMSE cutoff value of 24. Figure 3. D shows the distribution of diabetes, cardiovascular (CV) and nervous system (NS) medication usage among several participants based on ATC codes of medication configured in the application by caregivers.

### Principal Component Analysis

We performed a Principal Component Analysis (PCA) to group MMSE items into distinct components aimed at addressing cognitive dimensions in inpatients. PCA is particularly suited for this study due to its capacity to diminish data dimensionality while conserving the majority of variance. The PCA module of the Python sklearn.decomposition library was employed for this analysis. Figure 4. A demonstrates that 93% of the variance can be elucidated by the combined variations of principal components 1 and 2, suggesting that a 2D graph provides a robust approximation of the dataset. In Figure 4. B, where each dot represents the MMSE score of individual patients. In the plot patients with scores of ranges 12 and 22 cluster in lower values of PC1, while those between 27 and 30 cluster in higher values of both PC1 and PC2.

The eigenvectors in Figure 4. C indicate the direction in the feature space that captures the most variance in the data. Positive values suggest a positive correlation of each health variable with the principal component, while negative values suggest a negative correlation of that particular health variable with the principal component. Eigenvector percentages in 4.C along the first two Principal Components reveal that PC1 Steps and Move minutes positively contribute to this primary source of variation while HR and RHR exhibit distinct negative correlations. PC2 illustrates the negative correlation between sleep, RHR and the principal component. This spread distance between the PC1 patients and PC2 patients illustrates a very important phenomenon we observed in our results: patients exhibiting greater mobility demonstrate improved cardiovascular health, enhanced sleep quality, and are prominently clustered within the higher MMSE segment of the plot, indicative of better cognitive function.

### Correlation Coefficient Analysis

We plotted the correlation coefficient heatmap for all continuous smart wristband health variables in Figure 5 using Python stats.pearsonr package. Heatmap visualization shows that RHR is the most affecting variable targeting the cognitive scores.

### Chi-square Analysis

In addition to the analysis of non-categorical smart wristband-based health data variables, our dataset includes three categorical variables which we could not assess with PCA and correlation coefficient analysis: medication taken-or-not for glucose, medication taken-or-not for cardiovascular and medication taken-or-not for nervous system problems. For this, we computed the chi-square statistics between the smart wristband based health data variables using Python scipy.stats.chi2_contingency library and assessed the association between these three categorical variables with the cognitive scores.

Table 3 displays the chi-square results between taken medication for three disease groups with the MMSE scores. For this, we created 3 contingency tables with medication taken-or-not for glucose, cardiovascular and nervous system problems against MMSE scores. As expected, p- value of 0. 00498 along with a chi-square statistic of 7.8869 shows a strong association between the nervous system medication variable and MMSE score while other variables’ chi-square statistics were not significant.

**Table 3.** Chi-square values between patients using the three disease classed of medication and MMSE.

| **Nervous System Medication**<br>**vs MMSE Score** | **Diabetes Medication vs**<br>**MMSE Score** | **Cardiovascular**<br>**Medication vs MMSE**<br>**Score** |
| --- | --- | --- |
| Chi-square statistic: 7.8869<br>p-value: 0.00498<br>Degrees of freedom: 1 | Chi-square statistic: 3.5253<br>p-value: 0.06043<br>Degrees of freedom: 1 | Chi-square statistic: 0.56<br>p-value: 0.4551<br>Degrees of freedom: 1 |

The machine learning model we developed as a part of our previous study [12] was integrated into our mobile application to capture the predicted MMSE score of every participant at the beginning and end of the data collection period. At the beginning and end of the data collection period, each participant was requested to talk about the cookie theft picture used in the Boston Diagnostic Aphasia Exam (BDAE) for at most 5 minutes. Each participant’s voice description was assessed by our machine learning model and the model’s prediction score is used as the MMSE score of each participant. At the end of our study of 30 patients, 8 participants’ MMSE scores improved and 4 of these 8 participants had their RHRs dropped as much as 5% while increasing the average move minutes and steps by 24%. Figure 6 displays that the average increase in MMSE score was 9% for those with a final MMSE score of <= 24, whereas it was around 7% for those below 24. This result suggests that increased activity had a greater impact on individuals with lower MMSE scores. Subsequently, we calculated the correlation coefficient scores between the average number of steps, average RHR, and MMSE measured in the first and last weeks of data collection (see Table 4). The Pearson correlation coefficient between the change in RHR and the change in MMSE was found to be −0.48, with a p-value of 0.007. This indicates a significant inverse relationship between RHR and MMSE. The low p-value observed in the correlation between the change in average steps and RHR underscores the significance of physical activity in promoting improved cardiovascular health. Additionally, caregivers of these 8 participants reported enhanced mood and heightened awareness of well-being in their relatives, attributing these positive changes to the perceived health-tracking capabilities of smart wristbands.

**Figure 6.**
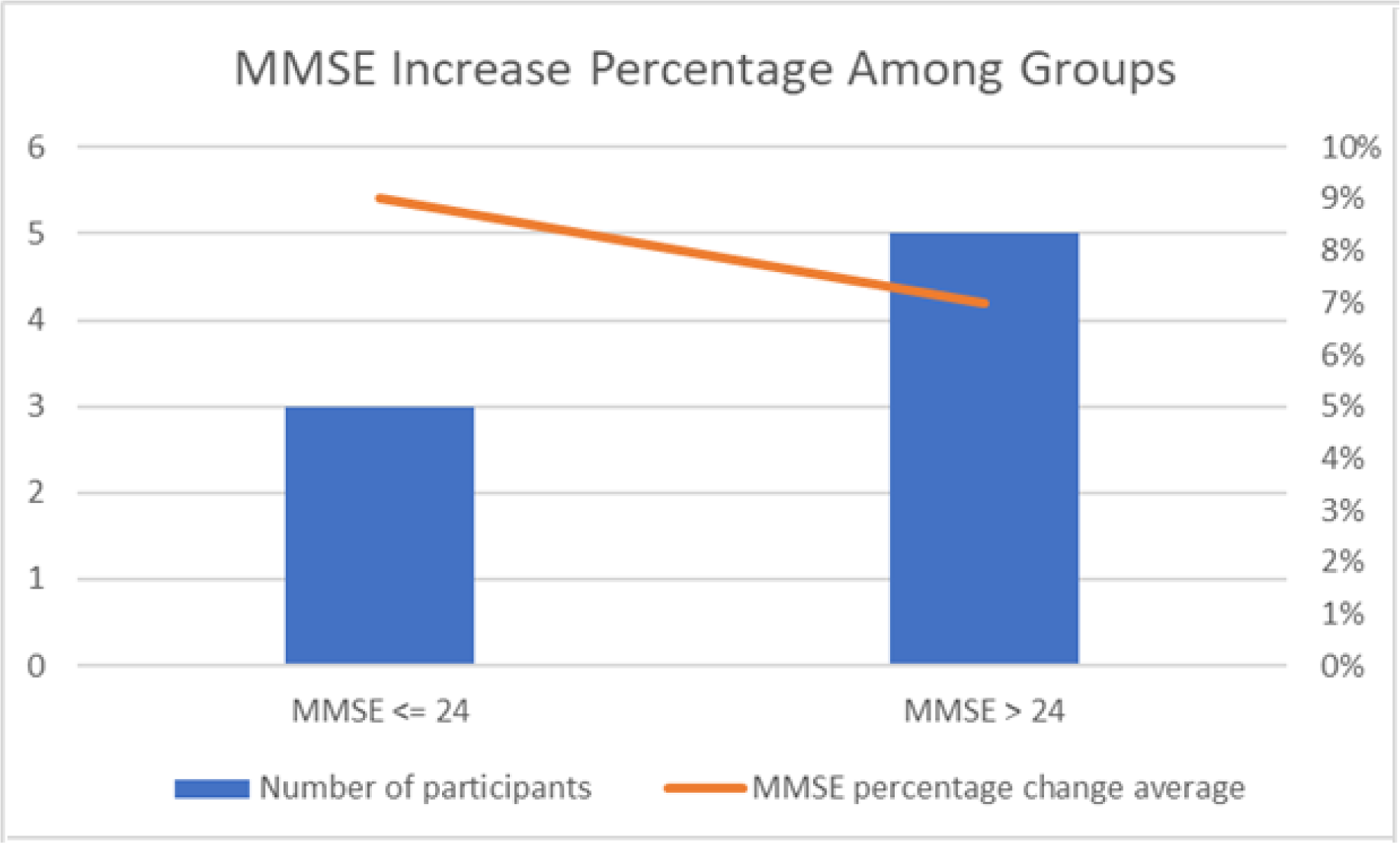
The rate of MMSE score increase among patients with lower and higher cognitive test scores.

**Table 4.** Pearson correlation coefficients indicating the strength of the correlation between changes in average steps, RHR, and MMSE.

|  |  |  |
| --- | --- | --- |
|  | Pearson Correlation Coefficient | p-value |
| **Average Steps change**<br>**vs MMSE change** | 0.41 | 0.03 |
| **RHR change vs MMSE**<br>**change** | -0.4288 | 0.007 |
| **Average Steps change**<br>**vs RHR change** | -90 | 4.88e-11 |

## Discussion

In our study involving 30 participants aged over 60, we demonstrated a significant improvement in cognitive test scores among patients with Mild Cognitive Impairment (MCI) attributed to increased physical activity. The cognitive decline in MCI is often subtle and does not significantly interfere with daily life, making it hard to distinguish from normal aging. The slight changes in memory and thinking skills are not easily captured by standard tests. Digital tests offer the advantage of continuous, non-invasive monitoring and behavioral insights, while blood tests provide biologically specific data through established clinical methods. Combining both approaches could potentially enhance the early detection and monitoring of MCI by leveraging the strengths of each method.

The primary innovation of our research lies in the introduction of a caregiver-oriented smart wristband-integrated software, enabling continuous monitoring of participants’ health parameters and lifestyle habits. Furthermore, our software analyzed both categorical and non-categorical features of MCI patient data for the first time, revealing that individuals with lower cognitive test scores exhibited approximately 50% less activity compared to those with higher scores. Another significant finding of our study is that our study unveiled those participants with higher cognitive test scores exhibited, on average, a 10% lower heart rate (HR) and resting heart rate (RHR) compared to those with lower scores, underscoring the pivotal role of heart health in cognitive decline.

8 patients who increased their activity by increasing their average number of steps and move minutes as much as 25% saw improvements in their MMSE tests that were conducted at the end of the study. We observed that these patients took wearing the smart wristbands as a challenge and continuously increased their activity which decreased their sleep time by 10% in average. Caregivers expressed increased positive mood among these 8 patients with a desire to continue using the smart wristband after the study.

The primary limitation of our study pertained to the recruitment of patients for the clinical trial. We encountered difficulties in persuading older patients, particularly those with MMSE scores below a certain threshold, to utilize smart wristbands, with a majority of these participants declining participation. Despite our concerted efforts to gather medication data from all participants, the incomplete nature of this data, particularly among the 16 patients with lower MMSE scores, necessitated the exclusion of this partial data from the statistical analysis.

Numerous earlier studies in the literature have linked health metrics such as Resting Heart Rate (RHR) with cognitive decline [3] [13] [14], a correlation our statistical analysis also corroborated by identifying RHR as the most influential factor for MMSE scores. Our findings suggest that continuous activity monitoring facilitated by devices like smart wristbands could serve as a pivotal strategy in monitoring and potentially attenuating the progression of cognitive decline.

## Data Availability

Anonymous patient health data is going to be available in the application database after acceptance

## Acknowledgment

We thank the Municipality of Izmir Dementia Clinic staff for their help in helping patients and caregivers in setting up mobile applications and smart wristbands and Dr. Barış Süzek for his invaluable contributions to the discussions about the method.

## Supporting Information

**S1 File. Ethics Committee Approval. Protocol Id : 230086**

**S2 File. Patient Consent Form**

**S3 File. Clinical Trial Record. Protocol Id: 230086**

## Notes

### Competing Interest Statement

The authors have declared no competing interest.

### Clinical Trial

NCT06417333

### Funding Statement

The author(s) received no specific funding for this work.

### Author Declarations

Non-interventional Ethics for Sports and Health Sciences- 2 Committee of Mugla Sitki Kocman University gave ethical approval for this work (protocol number:230086, October 18th, 2023).

